# Tomorrow’s physicians in distress: prevalence, socioeconomic gradients, and modifiable determinants of mental health problems among 1,560 medical students in Southern Brazil — a multicentre cross-sectional study

**DOI:** 10.64898/2026.07.14.26357843

**Authors:** Guilherme Noschang Vieira Bacchi, Laissa Harumi Furukawa, Alessandro Batista Soares, Aline Poli Turatti, Eduarda Gandolfi Horst, Gabriela Pinho Fillmann, Victoria Baptista dos Santos, Eduardo Karpovich, Vitória Pereira, Mariana Vieira Teles, Álvaro Righi de Leonço, Thales Fernando Canabarro Araujo, Kauane Scremin Reisdorfer, Marcela Menezes, Henrique Meneguetti, Isadora Castilho Firpo, Maria Isabel Martins Costa Kessler da Silveira, João Pedro Gonçalves Pacheco, Lucas Primo de Carvalho Alves, Bruno Luiz Guidolin, Jordana Tonezer Tedesco, Augusto Martins Lucas Bittencourt, Juliana Fernandes Tramontina, Catiane Zanin Cabral, Clara de Oliveira Lapa, Thiago Wendt Viola, Leonardo Araújo Pinto, Maria Dolores Braquehais Conesa, Lucas Spanemberg

## Abstract

**Background:** Medical students carry a disproportionate burden of mental health problems, but large multicentre studies from low- and middle-income countries remain scarce, and most evidence relies on single-institution samples and odds-ratio-based analyses that overstate associations for common outcomes. We provide a comprehensive epidemiological overview of mental health among medical students across an entire Brazilian state, quantifying the burden, its co-occurrence, and its socioeconomic and academic determinants.

**Methods:** Cross-sectional online survey of 1,560 medical students covering all 20 medical schools in operation in Rio Grande do Sul, Brazil (August–December 2023). Validated instruments assessed depressive and anxiety symptoms (PHQ-4), suicidal ideation (PHQ-9 item 9), non-suicidal self-injury, burnout (ESB-eu), quality of life (EUROHIS-QOL-8), spirituality (SSRS), substance use (ASSIST), prescription stimulant misuse, and mistreatment during training. Associations were estimated as adjusted prevalence ratios (aPR) using modified Poisson regression with robust variance, with linear trend tests, prespecified interactions, sensitivity analyses and E-values.

**Results:** Anxiety symptoms affected 64.5% (95% CI 62.1–66.9), depressive symptoms 45.5% (43.1–48.0), burnout 49.1% (46.7–51.6), recent suicidal ideation 20.6% (18.7–22.7), lifetime self-injury 13.1% (11.6–14.9) and stimulant misuse 7.9%; 57.4% screened positive for ≥2 outcomes. Quality of life declined monotonically with cumulative burden. Low family income showed inverse gradients across five outcomes (e.g., depression: PR 0.93 per income level, p = 0.002). Four factors were independently associated with nearly all outcomes: short sleep (<6 h; aPR up to 2.73), minority sexual orientation (aPR up to 2.02), family psychiatric history, and mistreatment during training (reported by 55.6%; aPR up to 1.49). Stimulant misuse tripled from the basic cycle to clerkship (aPR 2.30, 95% CI 1.36–3.88). There was no evidence of multiplicative interaction, indicating that the risk factors acted independently on the outcomes. Estimates were robust across sensitivity analyses.

**Conclusions:** One in two medical students in this state-wide sample screened positive for at least two mental health problems. Socioeconomic vulnerability, sleep deprivation, mistreatment and minority status operate as independent and largely modifiable determinants — actionable targets for medical schools and policymakers.

## Background

The mental health of medical students has become a global public health concern. Meta-analytic estimates indicate that approximately 27% of medical students worldwide present depressive symptoms and 11% report suicidal ideation [1], while pooled prevalences reach 34% for anxiety [2] and 37–44% for burnout before residency [3]. This burden exceeds that of age-matched peers in the general population and in most other undergraduate programmes [4–5], and carries consequences that extend beyond the individual: psychological distress during training predicts attrition, empathy erosion, unprofessional behaviour, and downstream physician burnout, with measurable effects on quality of care and patient safety [6–7]. Because today’s students constitute tomorrow’s workforce, their mental health is increasingly framed as a systems issue for health services rather than a private misfortune [7–8].

Three gaps limit the current evidence base. First, geography: most large studies originate from high-income countries, whereas low- and middle-income countries — where social adversity is more prevalent and mental health resources scarcer — remain underrepresented [9–11]. In Brazil, which operates the world’s second-largest system of medical schools [12], a national meta-analysis documented elevated prevalences of common mental disorders among medical students [13], but primary studies remain overwhelmingly restricted to single institutions or cities [14–15], precluding analyses of institutional and socioeconomic heterogeneity. Second, scope: depression, anxiety, burnout, suicidality, substance use, mistreatment and quality of life are usually studied in isolation, obscuring their co-occurrence within individuals — arguably the clinically decisive phenomenon. Third, methodology: most cross-sectional studies report odds ratios, which systematically overestimate associations when outcomes are common (prevalences of 20–65%, as is typical in this field), potentially distorting the perceived hierarchy of risk factors [16–17].

Beyond describing burden, identifying determinants that are modifiable at the institutional level is the priority for medical education. Converging evidence implicates socioeconomic disadvantage [10], sleep deprivation [18], mistreatment and discrimination during training [19], and minority sexual orientation [20] as candidate determinants, while the misuse of prescription stimulants as putative cognitive enhancers is an escalating concern in performance-driven academic cultures [21–22]. No Brazilian study has yet examined these domains simultaneously in a large multicentre sample.

We therefore conducted a state-wide, multicentre survey with complete institutional coverage: students from all 20 medical schools in operation in Rio Grande do Sul, Southern Brazil, were represented. We aimed to (i) estimate the prevalence and co-occurrence of depressive symptoms, anxiety symptoms, burnout, suicidal ideation, non-suicidal self-injury and stimulant misuse; (ii) quantify their socioeconomic and academic gradients; and (iii) identify independent, potentially modifiable determinants using prevalence-ratio-based models designed for common outcomes.

## Methods

### Design, setting and participants

This cross-sectional study is reported in accordance with the STROBE statement [23]. Between August and December 2023, medical students regularly enrolled in the medical schools of Rio Grande do Sul (RS), the southernmost Brazilian state (∼11 million inhabitants, 20 undergraduate medical programmes in operation at the time of data collection — 7 public and 13 private), were invited to complete a structured online questionnaire (Google Forms). Recruitment combined local coordinators at the seven co-participating institutions (PUCRS, UFN, UFSM, UNIJUÍ, FURG, UCPel and ULBRA) with open dissemination through institutional mailing lists, academic leagues, student associations and social media, allowing individual participation of students from all schools in the state. Participation was voluntary, anonymous and preceded by electronic informed consent; access was linked to a unique e-mail account to prevent multiple submissions, without investigator access to identifying information. All students aged ≥17 years enrolled in any of the six curricular years were eligible.

### Measures

The questionnaire comprised sociodemographic and academic variables (age, sex registered at birth, self-identified ethnicity, gender identity, sexual orientation, family income in Brazilian minimum wages [MW; R$ 1,320/month in 2023, approximately US$ 270 at the average exchange rate during data collection (US$ 1 ≈ R$ 4.90); the ≤4 MW and >20 MW strata therefore correspond to ≈ ≤US$ 1,080 and >US$ 5,400 per month], parental education, family medical and psychiatric history, institution and expected graduation year, from which the training stage was derived: basic cycle [years 1–2], intermediate cycle [years 3–4] and clerkship [years 5–6]), lifestyle variables (sleep duration, physical exercise, leisure activities), and the following validated instruments:

Depressive and anxiety symptoms: Patient Health Questionnaire-4 (PHQ-4), comprising the PHQ-2 and GAD-2 subscales; scores ≥3 on each subscale defined positive screening [24]. Suicidal ideation: PHQ-9 item 9 (thoughts of death or self-harm in the previous two weeks), analysed as any ideation (score ≥1) and recurrent ideation (frequency ≥2 episodes) [25]. Non-suicidal self-injury: lifetime self-reported behaviour aligned with DSM-5-TR criteria [26]. Burnout: Escala de Burnout de Estudantes (ESB-eu; 14 items, 0–4), with total score ≥20 defining positive screening [27]. Quality of life: EUROHIS-QOL-8 index [28–29]. Spirituality: Spirituality Self-Rating Scale (SSRS) [30–31]. Substance use: ASSIST-based items covering lifetime and past-three-month use, including electronic cigarettes [32]. Prescription stimulants: type (methylphenidate, lisdexamfetamine, modafinil), motives and access route; misuse was defined as use without prescription or beyond prescribed indications. Mistreatment: eight items on gender-based discrimination, humiliation, threats, physical harm and sexual harassment during medical school (0–3 frequency each); any exposure and total score were analysed.

Internal consistency in this sample ranged from α = 0.71 (mistreatment scale) to α = 0.92 (SSRS); all scores were computed from item-level responses (Supplementary Methods). The survey additionally included a measure of maladaptive personality traits (PID-5-BF+), whose results will be reported in a separate publication of this project.

### Data integrity

Of 1,576 submitted records, nine contained no data beyond the consent item or had consent declined, and seven duplicate participations (identified through matching voluntary contact information) were resolved by retaining the first submission, yielding an analytical sample of 1,560 students. A parsing defect in the exported file header was corrected before analysis, and all psychometric scores were recomputed from raw items rather than relying on platform-derived fields. UNIFRA and UFN (the same institution, renamed in 2018) were merged. Item-level missingness was <0.2% for all instruments; complete-case analysis was applied model-wise. A full data-integrity log is provided in the Supplementary Material.

### Statistical analysis

Prevalences are reported with 95% Wilson score confidence intervals. Bivariate comparisons used chi-square tests and t tests. Because all primary outcomes were common (8–65%), associations were estimated as prevalence ratios (PR) via modified Poisson regression with robust (sandwich) variance [16–17], avoiding the inflation of odds ratios in cross-sectional designs; logistic models were fitted in parallel for comparison. Two prespecified models were estimated for six outcomes (depressive symptoms, anxiety symptoms, burnout, suicidal ideation, self-injury, stimulant misuse): Model 1 (age, sex, family income in three strata, training stage, institution type) and Model 2 (Model 1 plus sexual orientation, short sleep [<6 h], family psychiatric history and mistreatment exposure). Income (five ordered levels) and training stage (three levels) were additionally modelled as continuous terms for linear trend tests. Prespecified multiplicative interactions (age × institution type; income × sex; cycle × sex; income × institution type) were tested. Robustness was examined by (i) excluding respondents with indeterminate or out-of-state institutions, (ii) retaining duplicate records, and (iii) E-values for unmeasured confounding [33]. Two-sided p < 0.05 defined significance. Analyses used Python 3.10; estimation routines were validated against reference implementations. Statistical programming was performed with the assistance of an artificial-intelligence tool under direct author specification and verification (see Declarations).

### Ethical considerations

The study was approved by the Research Ethics Committee of the Pontifical Catholic University of Rio Grande do Sul (CEP-PUCRS; consolidated approval report no. 5.784.468, issued 29 November 2022; CAAE 62723722.6.0000.5336), with a subsequently approved amendment including co-participating centres and local coordinators across the state (report no. 6.338.324, issued 2 October 2023), in accordance with Resolution 466/2012 of the Brazilian National Health Council and the Declaration of Helsinki. Appraisal by the national commission (CONEP) was not required. All participants provided electronic informed consent. Respondents could request individual feedback, and all received contact information for psychological support services.

## Results

### Sample characteristics

The final sample comprised 1,560 medical students from 20 medical schools (1091 [71.5%] private; 440 [28.5%] public). Mean age was 23.5 years (SD 3.7; range 17–44); 1129 (72.4%) were registered female at birth, 90.6% self-identified as White, 98.5% as cisgender and 21.2% reported a minority sexual orientation. Family income was ≤4 MW for 23.6% and >20 MW for 21.8%. Students were distributed across the basic (46.1%), intermediate (34.5%) and clerkship (19.4%) stages. A family psychiatric history was reported by 72.9%; 49.6% reported a personal psychiatric diagnosis and 13.1% current psychiatric medication (Table 1).

**Table 1.** Sociodemographic, academic and clinical characteristics (N = 1,560)

| Characteristic | n (%) / mean (SD) |
| --- | --- |
| Age, years — mean (SD) | 23.5 (3.7) |
| 17-20 years | 292 (18.7%) |
| 21-23 years | 633 (40.6%) |
| 24-26 years | 411 (26.3%) |
| ≥27 years | 224 (14.4%) |
| Female sex at birth | 1129 (72.4%) |
| White ethnicity | 1414 (90.6%) |
| Cisgender identity | 1537 (98.5%) |
| Minority sexual orientation | 328 (21.2%) |
| Family income ≤4 MW | 368 (23.6%) |
| Family income 4–20 MW | 852 (54.6%) |
| Family income >20 MW | 340 (21.8%) |
| Private institution | 1091 (71.3%) |
| Public institution | 440 (28.7%) |
| Basic cycle (years 1–2) | 719 (46.1%) |
| Intermediate cycle (years 3–4) | 538 (34.5%) |
| Clerkship (years 5–6) | 303 (19.4%) |
| Moved city for medical school | 992 (63.6%) |
| Physician in the family | 416 (26.7%) |
| Family psychiatric history | 1137 (72.9%) |
| Self-reported psychiatric diagnosis | 774 (49.6%) |
| Current psychiatric medication | 204 (13.1%) |
| Sleep hours — mean (SD) | 6.8 (1.1) |
| Short sleep (<6 h) | 128 (8.2%) |
| Regular physical exercise | 1204 (77.2%) |

MW: Brazilian minimum wage (R\$ 1,320/month in 2023; ~ US\$ 270). Income strata: ≤4 MW ≈ ≤US\$ 1,080/month; 4–20 MW ≈ US\$ 1,080–5,400; >20 MW ≈ >US\$ 5,400. Percentages over non-missing values.

### Overall burden and co-occurrence

Anxiety symptoms were present in 64.5% (95% CI 62.1–66.9; 1006/1559), depressive symptoms in 45.5% (95% CI 43.1–48.0; 710/1559), and both in 38.7% (95% CI 36.3–41.1; 603/1559). Burnout affected 49.1% (95% CI 46.7–51.6; 766/1559). Severe outcomes were frequent: 20.6% (95% CI 18.7–22.7; 321/1559) reported thoughts of death or self-harm in the previous two weeks, 6.9% (95% CI 5.8–8.3; 108/1559) recurrent ideation and 13.1% (95% CI 11.6–14.9; 205/1559) lifetime non-suicidal self-injury (Figure 1, Table 2).

**Figure 1.**
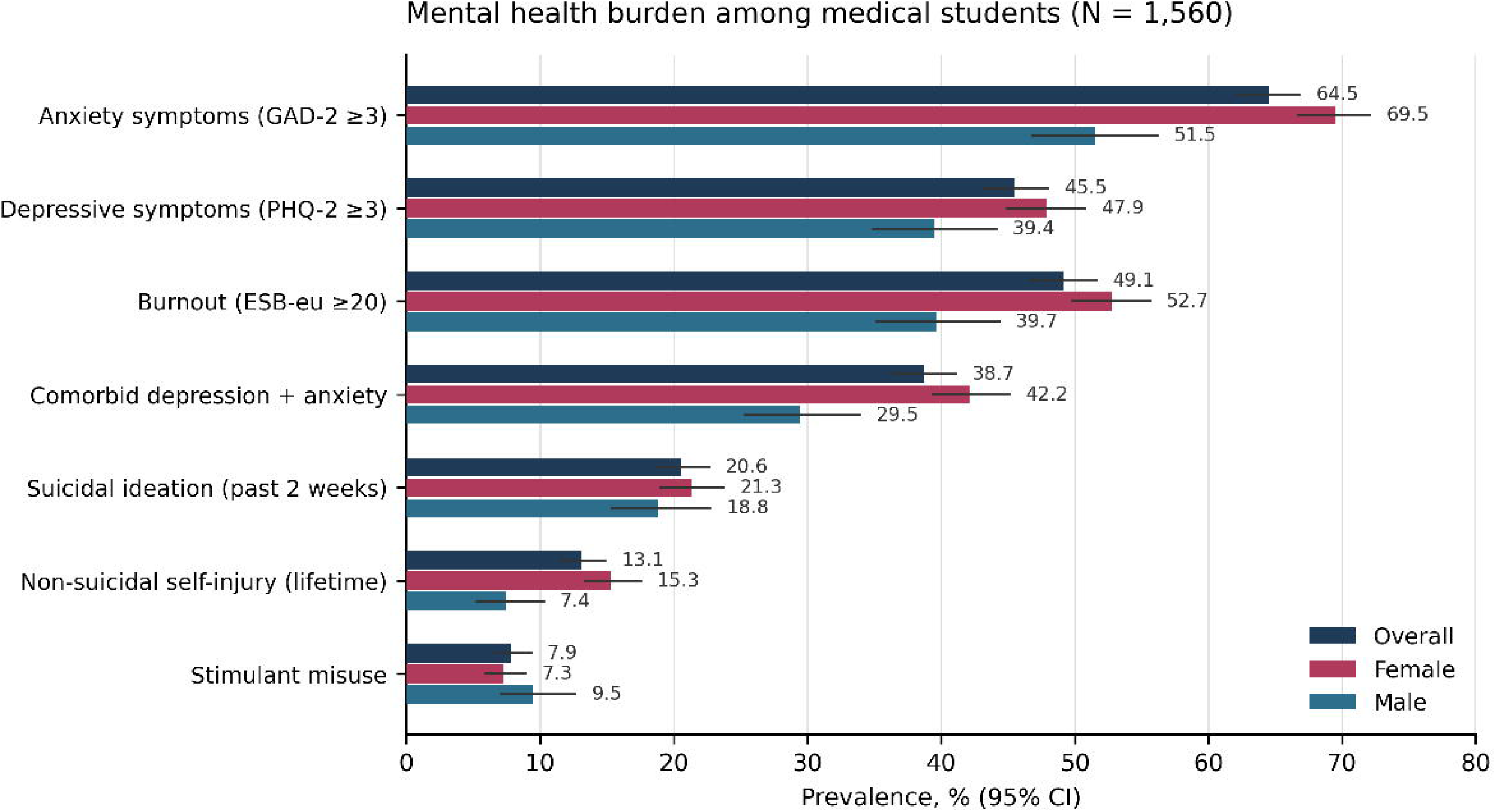
Prevalence (95% Wilson CI) of mental health outcomes among medical students in Rio Grande do Sul, Brazil (N = 1,560), overall and by sex at birth.

**Table 2.** Prevalence of outcomes, overall and by sex.

| Outcome | Overall % (95% CI) | Female % (n/N) | Male % (n/N) | p† |
| --- | --- | --- | --- | --- |
| Depressive symptoms (PHQ-2 ≥3) | 45.5 (43.1–48.0) | 47.9 (540/1128) | 39.4 (170/431) | 0.003 |
| Anxiety symptoms (GAD-2 ≥3) | 64.5 (62.1–66.9) | 69.5 (784/1128) | 51.5 (222/431) | <0.001 |
| Burnout (ESB-eu ≥20) | 49.1 (46.7–51.6) | 52.7 (595/1128) | 39.7 (171/431) | <0.001 |
| Suicidal ideation (2 weeks) | 20.6 (18.7–22.7) | 21.3 (240/1128) | 18.8 (81/431) | 0.278 |
| Recurrent ideation (≥2) | 6.9 (5.8–8.3) | 6.9 (78/1128) | 7.0 (30/431) | 0.975 |
| Self-injury (lifetime) | 13.1 (11.6–14.9) | 15.3 (173/1128) | 7.4 (32/431) | <0.001 |
| Stimulant use | 18.9 (17.1–20.9) | 18.4 (207/1128) | 20.4 (88/431) | 0.351 |
| Stimulant misuse | 7.9 (6.7–9.3) | 7.3 (82/1128) | 9.5 (41/431) | 0.142 |
| Mistreatment (any) | 55.6 (53.1–58.1) | 62.0 (699/1128) | 39.0 (168/431) | <0.001 |

† Chi-square test. CI: Wilson score interval.

Outcomes clustered within individuals: 57.2% screened positive for ≥2 of the five main outcomes and 36.4% for ≥3, while only 21.0% were negative on all five (Supplementary Figure S1). Recent suicidal ideation was nearly five times more frequent among students screening positive for depression (36.2% vs 7.5%). Quality of life decreased monotonically with cumulative burden, from 32.8 points (EUROHIS-QOL-8) among students with no positive screen to 23.8 among those with four or more (r = -0.59).

### Distribution across strata

Women showed higher prevalences of depressive symptoms (47.9% vs 39.4%; p = 0.003), anxiety symptoms (69.5% vs 51.5%; p < 0.001), burnout (52.7% vs 39.7%; p < 0.001) and lifetime self-injury (15.3% vs 7.4%; p < 0.001), whereas recent suicidal ideation did not differ by sex (p = 0.278). Mistreatment was reported by 62.0% of women and 39.0% of men (p < 0.001). Unadjusted income gradients were evident for depression, suicidal ideation and self-injury, and burnout peaked in the intermediate cycle while stimulant use and misuse rose toward clerkship (Figure 3; Supplementary Table S3).

**Figure 2.**
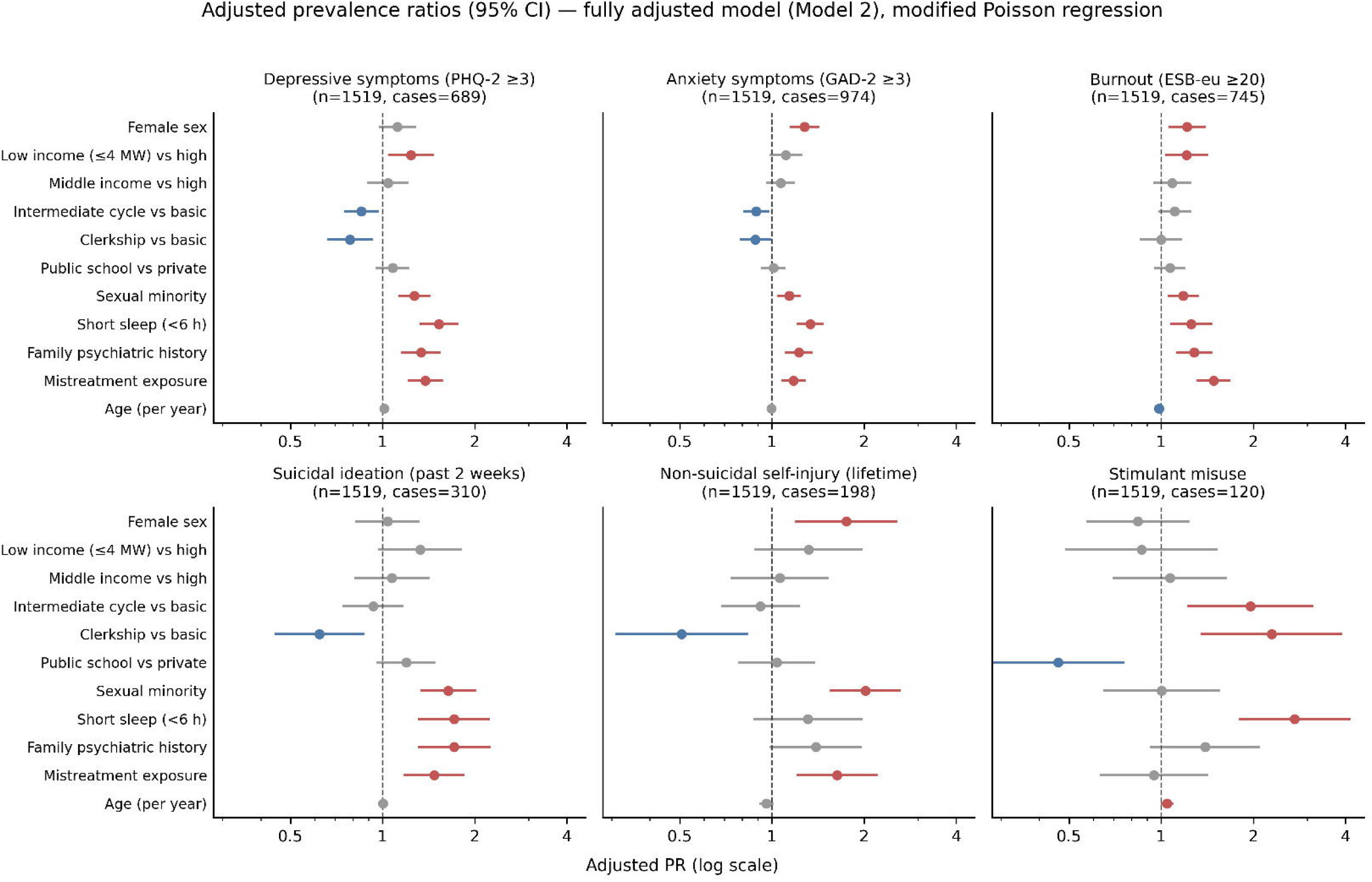
Adjusted prevalence ratios (95% CI) from fully adjusted modified Poisson models (Model 2) for six outcomes. Red: significant risk associations; blue: significant inverse associations; grey: non-significant.

**Figure 3.**
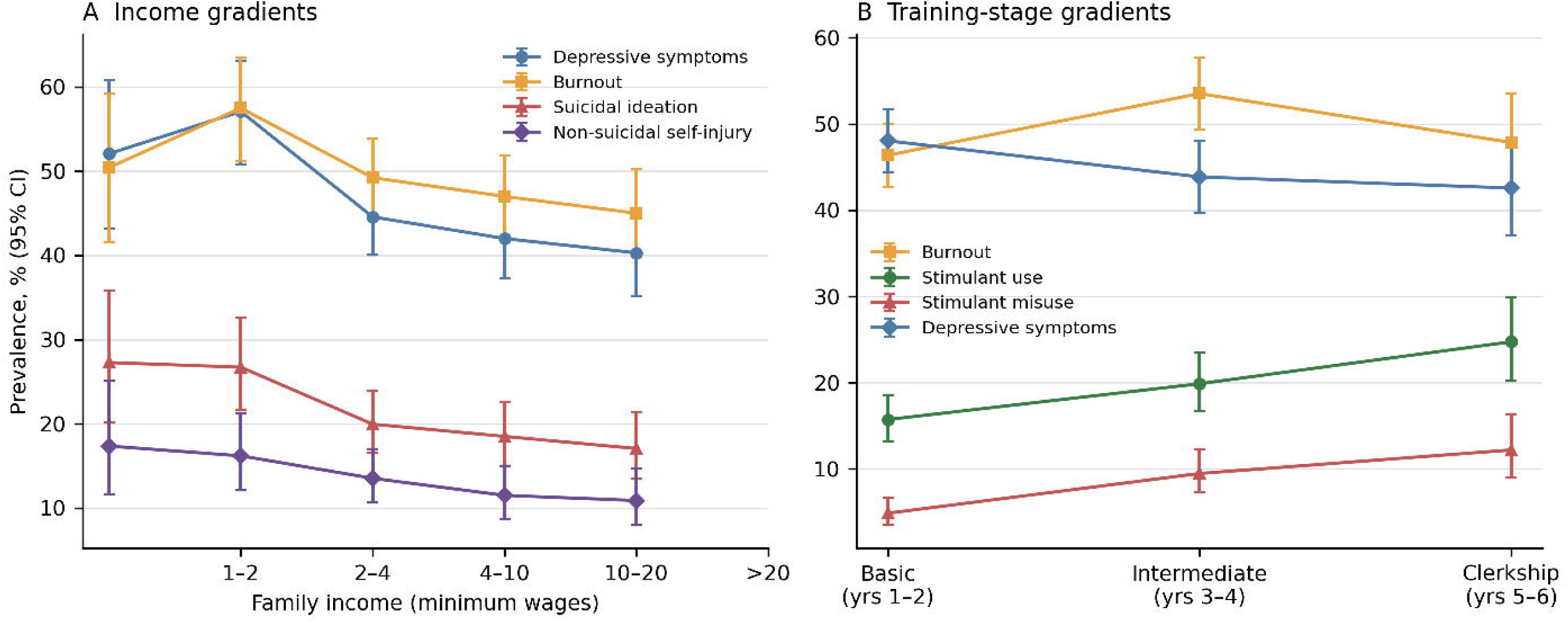
Unadjusted prevalence gradients (95% CI): (A) family income (five levels, minimum wages); (B) training stage.

### Adjusted determinants

In the core model (Model 1), female sex was independently associated with depressive symptoms (aPR 1.23 (95% CI 1.08–1.41)), anxiety symptoms (aPR 1.34 (95% CI 1.21–1.48)), burnout (aPR 1.36 (95% CI 1.19–1.55)) and self-injury (aPR 2.02 (95% CI 1.40–2.91)). Low family income (≤4 MW vs >20 MW) was associated with depressive symptoms (aPR 1.27 (95% CI 1.07–1.50)), anxiety symptoms (aPR 1.14 (95% CI 1.01–1.28)) and burnout (aPR 1.23 (95% CI 1.05–1.44)). Public-school enrolment was associated with suicidal ideation (aPR 1.31 (95% CI 1.06–1.63)) but with less than half the prevalence of stimulant misuse (aPR 0.45 (95% CI 0.27–0.75)) (Table 3, Figure 2).

**Table 3.**
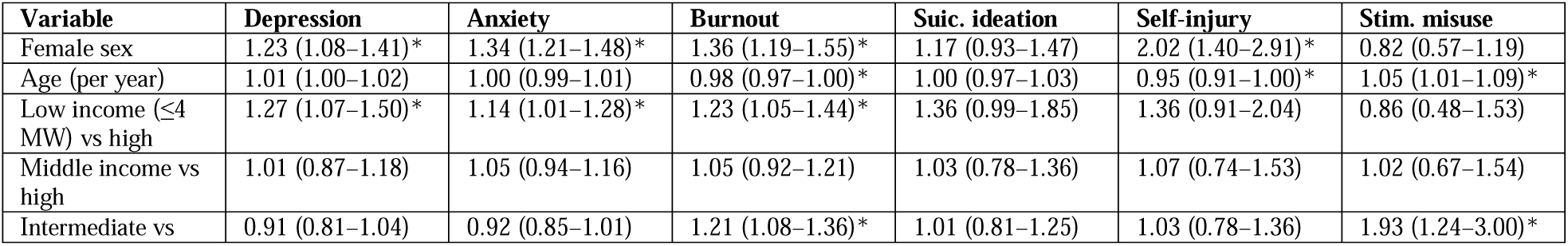
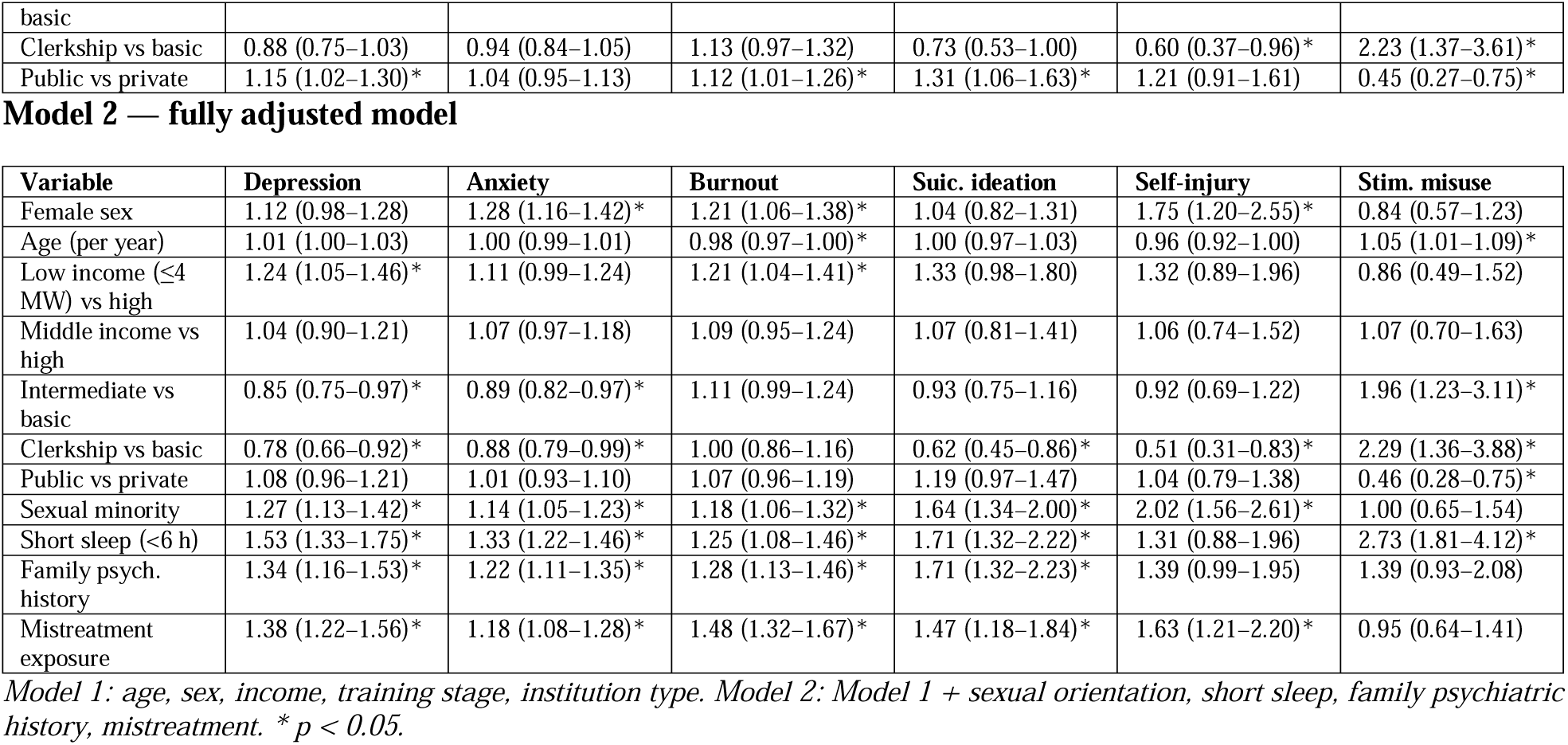
Adjusted prevalence ratios (95% CI) — modified Poisson regression Model 1 — core sociodemographic/academic model.

| Variable | Depression | Anxiety | Burnout | Suic. ideation | Self-injury | Stim. misuse |
| --- | --- | --- | --- | --- | --- | --- |
| Female sex | 1.23 (1.08–1.41)* | 1.34 (1.21–1.48)* | 1.36 (1.19–1.55)* | 1.17 (0.93–1.47) | 2.02 (1.40–2.91)* | 0.82 (0.57–1.19) |
| Age (per year) | 1.01 (1.00–1.02) | 1.00 (0.99–1.01) | 0.98 (0.97–1.00)* | 1.00 (0.97–1.03) | 0.95 (0.91–1.00)* | 1.05 (1.01–1.09)* |
| Low income (≤4 MW) vs high | 1.27 (1.07–1.50)* | 1.14 (1.01–1.28)* | 1.23 (1.05–1.44)* | 1.36 (0.99–1.85) | 1.36 (0.91–2.04) | 0.86 (0.48–1.53) |
| Middle income vs high | 1.01 (0.87–1.18) | 1.05 (0.94–1.16) | 1.05 (0.92–1.21) | 1.03 (0.78–1.36) | 1.07 (0.74–1.53) | 1.02 (0.67–1.54) |
| Intermediate vs | 0.91 (0.81–1.04) | 0.92 (0.85–1.01) | 1.21 (1.08–1.36)* | 1.01 (0.81–1.25) | 1.03 (0.78–1.36) | 1.93 (1.24–3.00)* |
| basic |  |  |  |  |  |  |
| Clerkship vs basic | 0.88 (0.75–1.03) | 0.94 (0.84–1.05) | 1.13 (0.97–1.32) | 0.73 (0.53–1.00) | 0.60 (0.37–0.96)* | 2.23 (1.37–3.61)* |
| Public vs private | 1.15 (1.02–1.30)* | 1.04 (0.95–1.13) | 1.12 (1.01–1.26)* | 1.31 (1.06–1.63)* | 1.21 (0.91–1.61) | 0.45 (0.27–0.75)* |

Model 2 — fully adjusted model
| Variable | Depression | Anxiety | Burnout | Suic. ideation | Self-injury | Stim. misuse |
| --- | --- | --- | --- | --- | --- | --- |
| Female sex | 1.12 (0.98–1.28) | 1.28 (1.16–1.42)* | 1.21 (1.06–1.38)* | 1.04 (0.82–1.31) | 1.75 (1.20–2.55)* | 0.84 (0.57–1.23) |
| Age (per year) | 1.01 (1.00–1.03) | 1.00 (0.99–1.01) | 0.98 (0.97–1.00)* | 1.00 (0.97–1.03) | 0.96 (0.92–1.00) | 1.05 (1.01–1.09)* |
| Low income (≤4 MW) vs high | 1.24 (1.05–1.46)* | 1.11 (0.99–1.24) | 1.21 (1.04–1.41)* | 1.33 (0.98–1.80) | 1.32 (0.89–1.96) | 0.86 (0.49–1.52) |
| Middle income vs high | 1.04 (0.90–1.21) | 1.07 (0.97–1.18) | 1.09 (0.95–1.24) | 1.07 (0.81–1.41) | 1.06 (0.74–1.52) | 1.07 (0.70–1.63) |
| Intermediate vs basic | 0.85 (0.75–0.97)* | 0.89 (0.82–0.97)* | 1.11 (0.99–1.24) | 0.93 (0.75–1.16) | 0.92 (0.69–1.22) | 1.96 (1.23–3.11)* |
| Clerkship vs basic | 0.78 (0.66–0.92)* | 0.88 (0.79–0.99)* | 1.00 (0.86–1.16) | 0.62 (0.45–0.86)* | 0.51 (0.31–0.83)* | 2.29 (1.36–3.88)* |
| Public vs private | 1.08 (0.96–1.21) | 1.01 (0.93–1.10) | 1.07 (0.96–1.19) | 1.19 (0.97–1.47) | 1.04 (0.79–1.38) | 0.46 (0.28–0.75)* |
| Sexual minority | 1.27 (1.13–1.42)* | 1.14 (1.05–1.23)* | 1.18 (1.06–1.32)* | 1.64 (1.34–2.00)* | 2.02 (1.56–2.61)* | 1.00 (0.65–1.54) |
| Short sleep (<6 h) | 1.53 (1.33–1.75)* | 1.33 (1.22–1.46)* | 1.25 (1.08–1.46)* | 1.71 (1.32–2.22)* | 1.31 (0.88–1.96) | 2.73 (1.81–4.12)* |
| Family psych. history | 1.34 (1.16–1.53)* | 1.22 (1.11–1.35)* | 1.28 (1.13–1.46)* | 1.71 (1.32–2.23)* | 1.39 (0.99–1.95) | 1.39 (0.93–2.08) |
| Mistreatment exposure | 1.38 (1.22–1.56)* | 1.18 (1.08–1.28)* | 1.48 (1.32–1.67)* | 1.47 (1.18–1.84)* | 1.63 (1.21–2.20)* | 0.95 (0.64–1.41) |

Model 1: age, sex, income, training stage, institution type. Model 2: Model 1 + sexual orientation, short sleep, family psychiatric history, mistreatment. \* $p < 0.05$ .

In the fully adjusted model (Model 2), four transversal factors were independently associated with nearly all outcomes: short sleep (aPR 1.53 for depression up to 2.73 for stimulant misuse), minority sexual orientation (up to aPR 2.02 for self-injury), family psychiatric history (aPR 1.71 (95% CI 1.32–2.23) for suicidal ideation) and mistreatment exposure (aPR 1.48 (95% CI 1.32–1.67) for burnout). The female-sex association with depression and anxiety attenuated after these adjustments — consistent with partial mediation through differential exposures — while the association with self-injury persisted (aPR 1.75 (95% CI 1.20–2.55)).

Trend tests confirmed inverse income gradients for depressive symptoms (PR 0.93 per level; p = 0.002), anxiety, burnout, suicidal ideation and self-injury, but not stimulant misuse, which showed a positive stage gradient (PR 1.49 per stage; p < 0.001). All prespecified interaction terms were non-significant (p > 0.42): there was no evidence of multiplicative interaction, indicating that the risk factors acted independently on the outcomes. Clerkship students showed lower prevalence of recent ideation (aPR 0.62 (95% CI 0.45–0.86)) and self-injury (aPR 0.51 (95% CI 0.31–0.83)) than basic-cycle students.

### Stimulants, substances and mistreatment

Prescription stimulants were used by 295 students (18.9%) — methylphenidate (n = 180), lisdexamfetamine (n = 106), modafinil (n = 9). Misuse was identified in 123 students (7.9% of the sample; 41.7% of users), with concentration, studying and staying awake as leading motives (Table 4). In adjusted models, misuse was associated with clerkship stage (aPR 2.29 (95% CI 1.36–3.88)), age, short sleep (aPR 2.73 (95% CI 1.81–4.12)) and private-school enrolment.

**Table 4.** Psychostimulant use and misuse (N = 1,560)

| Indicator | n (%) |
| --- | --- |
| Any prescription stimulant use | 295 (18.9%) |
| Methylphenidate | 180 |
| Lisdexamfetamine | 106 |
| Modafinil | 9 |
| Misuse | 123 (7.9% of sample; 41.7% of users) |
| Motives among users* |  |
| Improve concentration | 146 |
| Studying | 164 |
| Treat ADHD | 158 |
| Stay awake | 39 |
| Daytime sleepiness | 32 |
| Weight loss | 13 |
| Experimentation | 10 |
| Parties/recreational | 25 |

\* Multiple answers allowed; denominator = users ( $n = 295$ ).

Lifetime alcohol use was 94.5% (89.0% past three months; 34.3% at least weekly), electronic cigarettes 50.4%, cannabis 52.7%, tobacco 49.4% and amphetamines/ecstasy 20.2% (Supplementary Figure S2). Mistreatment during training was reported by 55.6% (95% CI 53.1–58.1; 867/1559) — most frequently sexist remarks (38.8%), public humiliation (28.8%) and unwanted sexual advances (20.7%) — and remained independently associated with depression, anxiety, burnout, suicidal ideation and self-injury (aPR 1.18–1.63; all p ≤ 0.001).

### Robustness

Estimates were essentially unchanged after excluding respondents with indeterminate/out-of-state institutions or retaining duplicates (maximum aPR change 0.04–0.05; Supplementary Table S5). Logistic models yielded identical inferences but systematically inflated estimates (e.g., OR 2.84 vs PR 1.53 for short sleep and depression; Supplementary Table S4, Figure S3). E-values ranged from 1.6 to 4.9 (Supplementary Table S8).

## Discussion

In this state-wide survey of 1,560 students representing all 20 medical schools in operation in Rio Grande do Sul — to our knowledge the first Brazilian study of medical student mental health with complete institutional coverage of an entire state — we found a burden that substantially exceeds global meta-analytic benchmarks: nearly half of students screened positive for depressive symptoms (45.5% vs ∼27% pooled globally [1]), almost two thirds for anxiety (64.5% vs ∼34% [2]), and one in five reported thoughts of death or self-harm in the preceding two weeks (20.6% vs ∼11% [1]). Burnout prevalence (49.1%) likewise exceeded pre-residency pooled estimates [3]. Three findings extend this descriptive picture in ways directly relevant to medical education policy.

First, co-occurrence is the rule, not the exception. More than half the sample (57.4%) screened positive for two or more outcomes, and quality of life declined monotonically with each additional problem — a dose-response pattern arguing that screening programmes limited to a single condition will systematically underestimate need. The near five-fold concentration of suicidal ideation among students with depressive symptoms reinforces recommendations to couple any depression screening with structured suicide-risk assessment [1, 8].

Second, socioeconomic position stratifies risk within an already privileged population. Even in medical schools — hardly a socioeconomically representative environment — we observed consistent inverse income gradients across five outcomes, robust to adjustment. This aligns with the social-determinants framework [10–11] and gains policy relevance in Brazil, where affirmative-action and student-financing policies have diversified the socioeconomic profile of medical cohorts [12]: widening access without proportional investment in student support risks converting educational inclusion into psychological attrition. The higher prevalence of suicidal ideation in public institutions, alongside markedly lower stimulant misuse, further suggests that institutional cultures shape distinct risk profiles rather than a single gradient of adversity.

Third, the principal determinants identified are modifiable. Short sleep — reported as <6 h/night by 8.7% — showed the largest and most pervasive associations (aPR 1.25–2.73), consistent with longitudinal evidence that sleep disturbance precedes and predicts mental disorder [18]. Mistreatment, reported by 55.6% (62.0% of women), matched the pooled 59.4% found in international training environments [19] and remained associated with all five clinical outcomes after full adjustment — the most consistent institution-level correlate we identified, and one for which schools bear direct responsibility. The doubled risk of self-injury among sexual-minority students replicates minority-stress findings in medical settings [20]. Notably, there was no evidence of multiplicative interaction, indicating that these factors acted independently on the outcomes: students combining low income, short sleep, minority status and mistreatment exposure therefore concentrate predictable, identifiable risk.

The stimulant findings warrant specific attention. Nearly one in five students used prescription stimulants and 7.9% met our misuse definition — 41.7% of users — with a steep gradient across training stages (aPR 2.30 for clerkship) and a strong association with short sleep, a pattern suggestive of pharmacological compensation for extended wakefulness in performance-driven contexts [21–22]. The halved prevalence in public schools points again to cultural and economic modulation of this behaviour.

The lower prevalence of recent suicidal ideation and lifetime self-injury among clerkship students deserves cautious interpretation. Rather than indicating that training becomes protective, this pattern is compatible with survivorship bias — students with severe symptoms interrupting or abandoning the course before clerkship — and with cohort effects, including pandemic-era entry classes in the basic cycle [14–15]. Longitudinal follow-up of this cohort, already planned within this project, will discriminate these hypotheses.

### Strengths and limitations

Strengths include the multicentre state-wide design with complete coverage of all 20 institutions (7 public, 13 private) in operation in 2023; the simultaneous assessment of symptoms, behaviours, lifestyle, mistreatment and substance use with validated instruments (α 0.71–0.92); a transparent data-integrity pipeline with recomputed scores, duplicate removal and documented corrections; and an estimation framework — prevalence ratios with robust variance, trend tests, interaction testing, sensitivity analyses and E-values — tailored to common outcomes and unmeasured confounding [16–17, 33]. Our direct comparison showing odds ratios inflating up to two-fold relative to prevalence ratios (Supplementary Figure S3) illustrates a methodological point with practical consequences for this literature.

Limitations must be acknowledged. First, the cross-sectional design precludes causal inference and temporal ordering; associations with mistreatment and sleep may be bidirectional. Second, convenience sampling without a verifiable sampling frame prevents response-rate calculation and may overrepresent distressed students (self-selection) or, conversely, exclude the most impaired; the female predominance (72.4%), although consistent with the feminisation of Brazilian medical cohorts [12], exceeds it. Third, institutional coverage was uneven — three schools contributed 42% of the sample — and 2.1% of respondents could not be assigned to an institution, although sensitivity analyses excluding them were unchanged. Fourth, all measures were self-reported, and PHQ-2/GAD-2 thresholds identify probable cases, not diagnoses; the ESB-eu cut-off, while internally consistent, lacks international calibration. Fifth, E-values of 1.6–4.9 indicate that moderate unmeasured confounding (e.g., pre-existing disorder severity) could account for the weaker associations. Finally, results from a single state with specific demographic characteristics may not generalise nationally, although RS hosts one of the country’s densest concentrations of medical schools [12].

### Implications

These data support a shift from awareness to accountability in medical education. Actionable implications include: (i) universal, multidimensional screening — brief instruments such as the PHQ-4 make simultaneous multi-outcome screening feasible at scale [24]; (ii) institutional monitoring of mistreatment with enforceable consequences, given its independent association with every clinical outcome examined [19, 34]; (iii) curricular protection of sleep, treating schedules incompatible with adequate rest as a patient-safety issue in formation [18, 35]; (iv) equity-oriented student support proportional to socioeconomic need, integrated with affirmative-action policies; and (v) explicit protocols addressing stimulant misuse in the clinical years. Because each risk factor was independently associated with the outcomes, simple counts of co-occurring exposures may be a pragmatic tool to stratify students for proactive outreach — a hypothesis to be tested formally in the longitudinal follow-up.

## Conclusions

In the largest multicentre assessment of medical student mental health conducted in Southern Brazil, one in two students screened positive for at least two mental health problems, with a burden well above global benchmarks. Socioeconomic vulnerability, short sleep, mistreatment during training, family psychiatric history and minority sexual orientation emerged as independent and largely modifiable determinants. Medical schools do not merely select vulnerable individuals; they administer environments whose characteristics — schedules, cultures, hierarchies and economic demands — are measurably associated with their students’ mental health. These findings provide a quantitative baseline for institutional intervention and for the longitudinal follow-up of this cohort.

## Supporting information

https://doi.org/10.6084/m9.figshare.32982245

## List of abbreviations

aPR: adjusted prevalence ratio
ASSIST: Alcohol, Smoking and Substance Involvement Screening Test
CI: confidence interval
ESB-eu: Escala de Burnout de Estudantes (university version)
GAD-2: Generalized Anxiety Disorder 2-item
MW: minimum wage
NSSI: non-suicidal self-injury
OR: odds ratio
PHQ: Patient Health Questionnaire
RS: Rio Grande do Sul
SSRS: Spirituality Self-Rating Scale
STROBE: Strengthening the Reporting of Observational Studies in Epidemiology.

## Declarations

### Ethics approval and consent to participate

The study was approved by the Research Ethics Committee of the Pontifical Catholic University of Rio Grande do Sul (CEP-PUCRS; approval report no. 5.784.468, 29 November 2022; amendment report no. 6.338.324, 2 October 2023; CAAE 62723722.6.0000.5336), in accordance with Resolution 466/2012 of the Brazilian National Health Council. All participants provided electronic informed consent before accessing the questionnaire.

### Consent for publication

Not applicable.

### Availability of data and materials

The de-identified dataset and analysis code are available from the corresponding author on reasonable request.

### Authors’ contributions

LS conceived and supervised all phases of the project and manuscript. GNVB, ABS, APT, EGH, GPF and VBS contributed to the initial study design, project writing, data collection and final revision. TWV contributed to data analysis and final revision. LHF, EK, VP, MVT, ARL, TC, KR, NB, MM, HM, HA, IF, MI, JPGP, LPCA, BLG, JTT, AMLB, JFT, KZC, COL, LAP and MDBC contributed to study dissemination, data collection and final revision. All authors read and approved the final manuscript.

## Data Availability

All data produced in this study are available from the authors upon reasonable request.

## Acknowledgements

The authors thank the participating students, the local coordinators at each medical school, and the academic leagues and student associations that supported recruitment.

## Use of artificial intelligence

During the preparation of this manuscript, the authors used Claude (Anthropic) as an artificial-intelligence assistant for: (i) statistical programming support, including data cleaning, implementation of statistical analyses pre-specified by the authors, and generation of figures, under the direct specification, verification and supervision of the authors; (ii) English translation and language editing, including grammar and spelling correction; and (iii) improvement of clarity, readability, and overall presentation of the manuscript. All study design decisions, statistical methods, analytical choices, results, interpretations, and final manuscript content were independently reviewed, verified, and approved by the authors, who take full responsibility for the accuracy and integrity of the work. No AI tool qualifies for authorship, in accordance with COPE recommendations and Springer Nature editorial policies.

