## Supplementary material for "Tomorrow’s physicians in distress: prevalence, socioeconomic gradients, and modifiable determinants of mental health problems among 1,560 medical students in Southern Brazil — a multicentre cross-sectional study": https://doi.org/10.6084/m9.figshare.32982245: Supplementary_Material_v21.docx

*Mental health burden among medical students in Southern Brazil: a multicentre epidemiological survey*

Contents: Supplementary Methods (data integrity log) · Tables S1–S8 · Figures S1–S3 · STROBE note

### Supplementary Methods — Data integrity and processing log

1. Source file: "Dados Escolas_Lucas.csv", 1,576 records × 359 variables, exported from the electronic survey platform (August–December 2023).

2. Header repair: the exported header contained an unescaped internal quotation mark in the PHQ-4 item 2 label ("Se sentir “para baixo”..."), which displaced the column mapping of all derived fields. The header was corrected before parsing; all analyses used the repaired file.

3. Exclusions: 9 records (0.6%) contained no data beyond the consent item (all submitted after the main collection window) or had consent declined; they were removed.

4. Duplicate participation: 7 respondents answered the questionnaire twice (identified by identical voluntary contact e-mails). The first submission of each was retained (primary analysis N = 1,560); a sensitivity analysis retaining duplicates is presented in Table S5.

5. Score reconstruction: all instrument scores (PHQ-4, EUROHIS-QOL-8, SSRS, ESB-eu, mistreatment scale) were recomputed from item-level responses. Agreement with spreadsheet-derived fields was 96–100%; discrepancies were traced to (a) the header defect above, (b) 3 records with spreadsheet formula errors in PHQ subscores, and (c) an inconsistent burnout threshold in the derived column (the operational cut-off consistent with the data is ESB-eu total ≥20). Recomputed scores were used throughout.

6. Institutions: 33 respondents (2.1%) reported an indeterminate institution or an institution outside Rio Grande do Sul; they were retained in primary analyses (their outcome data being valid) and excluded in sensitivity analysis (Table S5).

7. Item-level missingness was <0.2% for every instrument; complete-case analysis was applied model-wise.

### Table S1. Participating institutions

| **Institution** | **n** | **%** |
| --- | --- | --- |
| PUCRS - Porto Alegre | 377 | 24.2% |
| UFN/UNIFRA – Santa Maria | 146 | 9.4% |
| UFSM - Santa Maria | 145 | 9.3% |
| UCPEL - Pelotas | 137 | 8.8% |
| ULBRA - Canoas | 113 | 7.2% |
| UFPel - Pelotas | 109 | 7.0% |
| FURG - Rio Grande | 83 | 5.3% |
| UNIJUÍ - Ijuí | 81 | 5.2% |
| UNISINOS - São Leopoldo | 72 | 4.6% |
| UCS - Caxias do Sul | 65 | 4.2% |
| UFCSPA -Porto Alegre | 61 | 3.9% |
| UFRGS - Porto Alegre | 41 | 2.6% |
| URI - Erechim | 39 | 2.5% |
| Indefinido | 28 | 1.8% |
| IMED – Passo Fundo | 17 | 1.1% |
| UNISC - Universidade de Santa Cruz do Sul | 14 | 0.9% |
| FEEVALE - Novo Hamburgo | 11 | 0.7% |
| UPF - Passo Fundo | 7 | 0.4% |
| UNIVATES - Lajeado | 6 | 0.4% |
| UNIPAMPA - Uruguaiana | 3 | 0.2% |
| Uniara | 1 | 0.1% |
| Fam | 1 | 0.1% |
| Unc concórdia | 1 | 0.1% |
| UCB - DF | 1 | 0.1% |
| UFFS - Passo Fundo | 1 | 0.1% |

*Note: UNIFRA (Centro Universitário Franciscano) and UFN (Universidade Franciscana) refer to the same institution (renamed in 2018) and were merged. "Indeterminate" and out-of-state responses were retained in primary analyses and excluded in sensitivity analysis (Table S5).*

### Table S2. Self-reported psychiatric diagnoses (among the 49.6% reporting any diagnosis)

| **Diagnosis (self-reported)** | **n** | **% of total sample** |
| --- | --- | --- |
| Any anxiety disorder | 517 | 33.1% |
| Depression | 370 | 23.7% |
| ADHD | 90 | 5.8% |
| Bipolar disorder | 42 | 2.7% |
| Eating disorder | 98 | 6.3% |

*Note: multiple diagnoses allowed; categories aggregated from free-text and structured fields.*

### Table S3. Prevalence of outcomes across income levels and training stages (unadjusted, with 95% Wilson CI)

| **Outcome** | **Income** | **% (95% CI)** | **n/N** |
| --- | --- | --- | --- |
| Depression | 1–2 MW | 52.1 (43.2–60.8) | 63/121 |
|  | 2–4 MW | 57.1 (50.9–63.1) | 141/247 |
|  | 4–10 MW | 44.6 (40.0–49.2) | 201/451 |
|  | 10–20 MW | 42.0 (37.3–46.9) | 168/400 |
|  | >20 MW | 40.3 (35.2–45.6) | 137/340 |
| Anxiety | 1–2 MW | 67.8 (59.0–75.4) | 82/121 |
|  | 2–4 MW | 70.9 (64.9–76.2) | 175/247 |
|  | 4–10 MW | 64.3 (59.8–68.6) | 290/451 |
|  | 10–20 MW | 64.2 (59.4–68.8) | 257/400 |
|  | >20 MW | 59.4 (54.1–64.5) | 202/340 |
| Burnout | 1–2 MW | 50.4 (41.6–59.2) | 61/121 |
|  | 2–4 MW | 57.5 (51.3–63.5) | 142/247 |
|  | 4–10 MW | 49.2 (44.6–53.8) | 222/451 |
|  | 10–20 MW | 47.0 (42.2–51.9) | 188/400 |
|  | >20 MW | 45.0 (39.8–50.3) | 153/340 |
| Suicidal ideation | 1–2 MW | 27.3 (20.1–35.8) | 33/121 |
|  | 2–4 MW | 26.7 (21.6–32.6) | 66/247 |
|  | 4–10 MW | 20.0 (16.5–23.9) | 90/451 |
|  | 10–20 MW | 18.5 (15.0–22.6) | 74/400 |
|  | >20 MW | 17.1 (13.4–21.4) | 58/340 |
| Self-injury | 1–2 MW | 17.4 (11.6–25.1) | 21/121 |
|  | 2–4 MW | 16.2 (12.1–21.3) | 40/247 |
|  | 4–10 MW | 13.5 (10.7–17.0) | 61/451 |
|  | 10–20 MW | 11.5 (8.7–15.0) | 46/400 |
|  | >20 MW | 10.9 (8.0–14.6) | 37/340 |
| Stimulant misuse | 1–2 MW | 6.6 (3.4–12.5) | 8/121 |
|  | 2–4 MW | 5.3 (3.1–8.8) | 13/247 |
|  | 4–10 MW | 7.8 (5.6–10.6) | 35/451 |
|  | 10–20 MW | 8.5 (6.1–11.6) | 34/400 |
|  | >20 MW | 9.7 (7.0–13.3) | 33/340 |

| **Outcome** | **Stage** | **% (95% CI)** | **n/N** |
| --- | --- | --- | --- |
| Depression | Basic | 48.1 (44.4–51.7) | 345/718 |
|  | Intermediate | 43.9 (39.7–48.1) | 236/538 |
|  | Clerkship | 42.6 (37.1–48.2) | 129/303 |
| Anxiety | Basic | 67.7 (64.2–71.0) | 486/718 |
|  | Intermediate | 61.5 (57.3–65.5) | 331/538 |
|  | Clerkship | 62.4 (56.8–67.6) | 189/303 |
| Burnout | Basic | 46.4 (42.8–50.0) | 333/718 |
|  | Intermediate | 53.5 (49.3–57.7) | 288/538 |
|  | Clerkship | 47.9 (42.3–53.5) | 145/303 |
| Suicidal ideation | Basic | 21.9 (19.0–25.0) | 157/718 |
|  | Intermediate | 21.7 (18.5–25.4) | 117/538 |
|  | Clerkship | 15.5 (11.9–20.0) | 47/303 |
| Self-injury | Basic | 15.2 (12.7–18.0) | 109/718 |
|  | Intermediate | 13.8 (11.1–16.9) | 74/538 |
|  | Clerkship | 7.3 (4.8–10.7) | 22/303 |
| Stimulant misuse | Basic | 4.9 (3.5–6.7) | 35/718 |
|  | Intermediate | 9.5 (7.3–12.3) | 51/538 |
|  | Clerkship | 12.2 (9.0–16.4) | 37/303 |

### Table S4. Comparison of adjusted prevalence ratios (modified Poisson) and adjusted odds ratios (logistic) — fully adjusted models

| **Outcome** | **Variable** | **aPR (95% CI)** | **aOR (95% CI)** |
| --- | --- | --- | --- |
| Depression | Female sex | 1.12 (0.98–1.28) | 1.25 (0.97–1.61) |
|  | Low income vs high | 1.24 (1.05–1.46) | 1.54 (1.11–2.15) |
|  | Clerkship vs basic | 0.78 (0.66–0.92) | 0.63 (0.46–0.86) |
|  | Public school | 1.08 (0.96–1.21) | 1.17 (0.92–1.50) |
|  | Sexual minority | 1.27 (1.13–1.42) | 1.68 (1.29–2.19) |
|  | Short sleep | 1.53 (1.33–1.75) | 2.82 (1.86–4.28) |
|  | Family psych. history | 1.34 (1.16–1.53) | 1.68 (1.31–2.15) |
|  | Mistreatment | 1.38 (1.22–1.56) | 1.83 (1.45–2.30) |
| Anxiety | Female sex | 1.28 (1.16–1.42) | 1.92 (1.50–2.47) |
|  | Low income vs high | 1.11 (0.99–1.24) | 1.35 (0.96–1.92) |
|  | Clerkship vs basic | 0.88 (0.79–0.99) | 0.70 (0.50–0.97) |
|  | Public school | 1.01 (0.93–1.10) | 1.04 (0.80–1.35) |
|  | Sexual minority | 1.14 (1.05–1.23) | 1.56 (1.17–2.08) |
|  | Short sleep | 1.33 (1.22–1.46) | 3.20 (1.92–5.33) |
|  | Family psych. history | 1.22 (1.11–1.35) | 1.73 (1.36–2.20) |
|  | Mistreatment | 1.18 (1.08–1.28) | 1.58 (1.25–1.99) |
| Burnout | Female sex | 1.21 (1.06–1.38) | 1.46 (1.14–1.88) |
|  | Low income vs high | 1.21 (1.04–1.41) | 1.50 (1.08–2.09) |
|  | Clerkship vs basic | 1.00 (0.86–1.16) | 1.00 (0.73–1.37) |
|  | Public school | 1.07 (0.96–1.19) | 1.16 (0.91–1.49) |
|  | Sexual minority | 1.18 (1.06–1.32) | 1.49 (1.14–1.94) |
|  | Short sleep | 1.25 (1.08–1.46) | 1.68 (1.13–2.51) |
|  | Family psych. history | 1.28 (1.13–1.46) | 1.63 (1.28–2.07) |
|  | Mistreatment | 1.48 (1.32–1.67) | 2.17 (1.73–2.72) |
| Suicidal ideation | Female sex | 1.04 (0.82–1.31) | 1.06 (0.77–1.44) |
|  | Low income vs high | 1.33 (0.98–1.80) | 1.46 (0.97–2.18) |
|  | Clerkship vs basic | 0.62 (0.45–0.86) | 0.54 (0.36–0.82) |
|  | Public school | 1.19 (0.97–1.47) | 1.27 (0.95–1.70) |
|  | Sexual minority | 1.64 (1.34–2.00) | 1.98 (1.49–2.65) |
|  | Short sleep | 1.71 (1.32–2.22) | 2.15 (1.42–3.27) |
|  | Family psych. history | 1.71 (1.32–2.23) | 1.96 (1.41–2.74) |
|  | Mistreatment | 1.47 (1.18–1.84) | 1.65 (1.24–2.20) |
| Self-injury | Female sex | 1.75 (1.20–2.55) | 1.93 (1.27–2.94) |
|  | Low income vs high | 1.32 (0.89–1.96) | 1.40 (0.87–2.27) |
|  | Clerkship vs basic | 0.51 (0.31–0.83) | 0.45 (0.26–0.78) |
|  | Public school | 1.04 (0.79–1.38) | 1.05 (0.74–1.50) |
|  | Sexual minority | 2.02 (1.56–2.61) | 2.40 (1.72–3.36) |
|  | Short sleep | 1.31 (0.88–1.96) | 1.41 (0.83–2.39) |
|  | Family psych. history | 1.39 (0.99–1.95) | 1.47 (1.00–2.18) |
|  | Mistreatment | 1.63 (1.21–2.20) | 1.79 (1.27–2.54) |
| Stimulant misuse | Female sex | 0.84 (0.57–1.23) | 0.81 (0.52–1.26) |
|  | Low income vs high | 0.86 (0.49–1.52) | 0.84 (0.45–1.56) |
|  | Clerkship vs basic | 2.29 (1.36–3.88) | 2.50 (1.46–4.31) |
|  | Public school | 0.46 (0.28–0.75) | 0.42 (0.25–0.72) |
|  | Sexual minority | 1.00 (0.65–1.54) | 1.01 (0.63–1.63) |
|  | Short sleep | 2.73 (1.81–4.12) | 3.30 (1.96–5.56) |
|  | Family psych. history | 1.39 (0.93–2.08) | 1.45 (0.91–2.30) |
|  | Mistreatment | 0.95 (0.64–1.41) | 0.95 (0.62–1.44) |

### Table S5. Sensitivity analyses — fully adjusted prevalence ratios

| **Outcome** | **Variable** | **Primary aPR** | **Excl. indeterminate institutions** | **Retaining duplicates** |
| --- | --- | --- | --- | --- |
| Depression | Female sex | 1.12 | 1.11 (0.98–1.27) | 1.12 (0.98–1.28) |
|  | Low income vs high | 1.24 | 1.23 (1.05–1.45) | 1.23 (1.05–1.45) |
|  | Clerkship vs basic | 0.78 | 0.79 (0.67–0.93) | 0.78 (0.66–0.92) |
|  | Public school | 1.08 | 1.08 (0.96–1.21) | 1.09 (0.97–1.22) |
|  | Sexual minority | 1.27 | 1.27 (1.13–1.42) | 1.26 (1.13–1.42) |
|  | Short sleep | 1.53 | 1.53 (1.33–1.75) | 1.53 (1.34–1.75) |
|  | Family psych. history | 1.34 | 1.34 (1.17–1.54) | 1.33 (1.16–1.53) |
|  | Mistreatment | 1.38 | 1.38 (1.22–1.57) | 1.38 (1.22–1.56) |
| Anxiety | Female sex | 1.28 | 1.27 (1.15–1.41) | 1.28 (1.15–1.41) |
|  | Low income vs high | 1.11 | 1.11 (0.99–1.24) | 1.11 (1.00–1.25) |
|  | Clerkship vs basic | 0.88 | 0.89 (0.79–0.99) | 0.89 (0.80–0.99) |
|  | Public school | 1.01 | 1.01 (0.93–1.10) | 1.01 (0.93–1.10) |
|  | Sexual minority | 1.14 | 1.14 (1.05–1.23) | 1.14 (1.05–1.24) |
|  | Short sleep | 1.33 | 1.33 (1.22–1.46) | 1.33 (1.22–1.46) |
|  | Family psych. history | 1.22 | 1.23 (1.12–1.35) | 1.22 (1.11–1.35) |
|  | Mistreatment | 1.18 | 1.18 (1.09–1.28) | 1.18 (1.08–1.27) |
| Burnout | Female sex | 1.21 | 1.22 (1.07–1.39) | 1.22 (1.07–1.39) |
|  | Low income vs high | 1.21 | 1.21 (1.04–1.42) | 1.20 (1.03–1.39) |
|  | Clerkship vs basic | 1.00 | 1.00 (0.86–1.16) | 1.00 (0.86–1.16) |
|  | Public school | 1.07 | 1.07 (0.96–1.19) | 1.08 (0.97–1.20) |
|  | Sexual minority | 1.18 | 1.18 (1.06–1.32) | 1.18 (1.06–1.32) |
|  | Short sleep | 1.25 | 1.26 (1.08–1.46) | 1.25 (1.08–1.45) |
|  | Family psych. history | 1.28 | 1.28 (1.12–1.46) | 1.28 (1.13–1.46) |
|  | Mistreatment | 1.48 | 1.48 (1.31–1.67) | 1.48 (1.31–1.66) |
| Suicidal ideation | Female sex | 1.04 | 1.04 (0.82–1.31) | 1.04 (0.82–1.31) |
|  | Low income vs high | 1.33 | 1.33 (0.98–1.80) | 1.32 (0.97–1.79) |
|  | Clerkship vs basic | 0.62 | 0.62 (0.45–0.86) | 0.62 (0.45–0.86) |
|  | Public school | 1.19 | 1.19 (0.96–1.47) | 1.20 (0.97–1.48) |
|  | Sexual minority | 1.64 | 1.63 (1.34–2.00) | 1.63 (1.34–2.00) |
|  | Short sleep | 1.71 | 1.71 (1.31–2.22) | 1.72 (1.32–2.24) |
|  | Family psych. history | 1.71 | 1.71 (1.32–2.23) | 1.71 (1.31–2.22) |
|  | Mistreatment | 1.47 | 1.48 (1.18–1.84) | 1.49 (1.19–1.85) |
| Self-injury | Female sex | 1.75 | 1.73 (1.19–2.52) | 1.80 (1.23–2.62) |
|  | Low income vs high | 1.32 | 1.32 (0.88–1.96) | 1.32 (0.89–1.95) |
|  | Clerkship vs basic | 0.51 | 0.51 (0.31–0.83) | 0.52 (0.32–0.85) |
|  | Public school | 1.04 | 1.04 (0.79–1.38) | 1.05 (0.79–1.38) |
|  | Sexual minority | 2.02 | 2.02 (1.56–2.62) | 1.98 (1.53–2.56) |
|  | Short sleep | 1.31 | 1.32 (0.88–1.97) | 1.30 (0.87–1.94) |
|  | Family psych. history | 1.39 | 1.43 (1.01–2.01) | 1.37 (0.98–1.91) |
|  | Mistreatment | 1.63 | 1.66 (1.23–2.23) | 1.63 (1.22–2.18) |
| Stimulant misuse | Female sex | 0.84 | 0.84 (0.57–1.22) | 0.84 (0.57–1.23) |
|  | Low income vs high | 0.86 | 0.86 (0.49–1.51) | 0.86 (0.49–1.50) |
|  | Clerkship vs basic | 2.29 | 2.30 (1.36–3.88) | 2.29 (1.36–3.87) |
|  | Public school | 0.46 | 0.46 (0.28–0.75) | 0.46 (0.28–0.75) |
|  | Sexual minority | 1.00 | 1.00 (0.65–1.54) | 1.00 (0.65–1.53) |
|  | Short sleep | 2.73 | 2.72 (1.80–4.11) | 2.74 (1.81–4.14) |
|  | Family psych. history | 1.39 | 1.39 (0.93–2.09) | 1.39 (0.93–2.08) |
|  | Mistreatment | 0.95 | 0.95 (0.64–1.42) | 0.95 (0.64–1.42) |

### Table S6. Linear trend tests (adjusted for age, sex and institution type)

| **Outcome** | **Trend term** | **PR per level (95% CI)** | **p** |
| --- | --- | --- | --- |
| Depression | Income (per level increase) | 0.928 (0.885–0.972) | 0.002 |
| Depression | Cycle (per stage) | 0.918 (0.850–0.992) | 0.030 |
| Anxiety | Income (per level increase) | 0.965 (0.935–0.996) | 0.027 |
| Anxiety | Cycle (per stage) | 0.953 (0.904–1.004) | 0.071 |
| Burnout | Income (per level increase) | 0.955 (0.914–0.998) | 0.039 |
| Burnout | Cycle (per stage) | 1.071 (0.998–1.148) | 0.057 |
| Suicidal ideation | Income (per level increase) | 0.905 (0.829–0.987) | 0.024 |
| Suicidal ideation | Cycle (per stage) | 0.867 (0.755–0.996) | 0.044 |
| Self-injury | Income (per level increase) | 0.889 (0.795–0.995) | 0.040 |
| Self-injury | Cycle (per stage) | 0.819 (0.678–0.989) | 0.038 |
| Stimulant misuse | Income (per level increase) | 1.080 (0.925–1.260) | 0.330 |
| Stimulant misuse | Cycle (per stage) | 1.495 (1.206–1.854) | <0.001 |

### Table S7. Instrument scores and internal consistency

| **Instrument** | **Cronbach α** | **Mean ± SD** |
| --- | --- | --- |
| PHQ-4 (0–12) | 0.81 | 6.03 ± 3.08 |
| EUROHIS-QOL-8 (8–40) | 0.84 | 28.4 ± 5.4 |
| SSRS spirituality (6–30) | 0.92 | 15.49 ± 7.09 |
| ESB-eu burnout (0–56) | 0.89 | 20.24 ± 9.84 |
| Mistreatment scale (0–24) | 0.71 | 1.86 ± 2.64 |

### Table S8. E-values for significant fully adjusted associations

| **Outcome** | **Variable** | **aPR** | **E-value (point)** | **E-value (CI bound)** |
| --- | --- | --- | --- | --- |
| Depression | Low income | 1.24 | 1.78 | 1.29 |
| Depression | Intermediate cycle | 0.85 | 1.62 | 1.23 |
| Depression | Clerkship | 0.78 | 1.87 | 1.38 |
| Depression | Sexual minority | 1.27 | 1.85 | 1.52 |
| Depression | Short sleep | 1.53 | 2.42 | 2.00 |
| Depression | Family psych. history | 1.34 | 2.00 | 1.60 |
| Depression | Mistreatment | 1.38 | 2.10 | 1.74 |
| Anxiety | Female sex | 1.28 | 1.88 | 1.58 |
| Anxiety | Intermediate cycle | 0.89 | 1.50 | 1.21 |
| Anxiety | Clerkship | 0.88 | 1.52 | 1.13 |
| Anxiety | Sexual minority | 1.14 | 1.53 | 1.28 |
| Anxiety | Short sleep | 1.33 | 2.00 | 1.73 |
| Anxiety | Family psych. history | 1.22 | 1.75 | 1.47 |
| Anxiety | Mistreatment | 1.18 | 1.63 | 1.39 |
| Burnout | Age (per year) | 0.98 | 1.14 | 1.01 |
| Burnout | Female sex | 1.21 | 1.72 | 1.33 |
| Burnout | Low income | 1.21 | 1.72 | 1.24 |
| Burnout | Sexual minority | 1.18 | 1.64 | 1.31 |
| Burnout | Short sleep | 1.25 | 1.82 | 1.38 |
| Burnout | Family psych. history | 1.28 | 1.89 | 1.51 |
| Burnout | Mistreatment | 1.48 | 2.33 | 1.97 |
| Suicidal ideation | Clerkship | 0.62 | 2.60 | 1.59 |
| Suicidal ideation | Sexual minority | 1.64 | 2.66 | 2.02 |
| Suicidal ideation | Short sleep | 1.71 | 2.81 | 1.96 |
| Suicidal ideation | Family psych. history | 1.71 | 2.82 | 1.96 |
| Suicidal ideation | Mistreatment | 1.47 | 2.31 | 1.65 |
| Self-injury | Female sex | 1.75 | 2.89 | 1.70 |
| Self-injury | Clerkship | 0.51 | 3.35 | 1.71 |
| Self-injury | Sexual minority | 2.02 | 3.45 | 2.49 |
| Self-injury | Mistreatment | 1.63 | 2.65 | 1.73 |
| Stimulant misuse | Age (per year) | 1.05 | 1.26 | 1.09 |
| Stimulant misuse | Intermediate cycle | 1.96 | 3.32 | 1.76 |
| Stimulant misuse | Clerkship | 2.29 | 4.02 | 2.06 |
| Stimulant misuse | Public school | 0.46 | 3.76 | 1.99 |
| Stimulant misuse | Short sleep | 2.73 | 4.90 | 3.01 |

*E-value: minimum strength of association (risk-ratio scale) that an unmeasured confounder would need with both exposure and outcome to fully explain the observed association.*

### Supplementary Figures

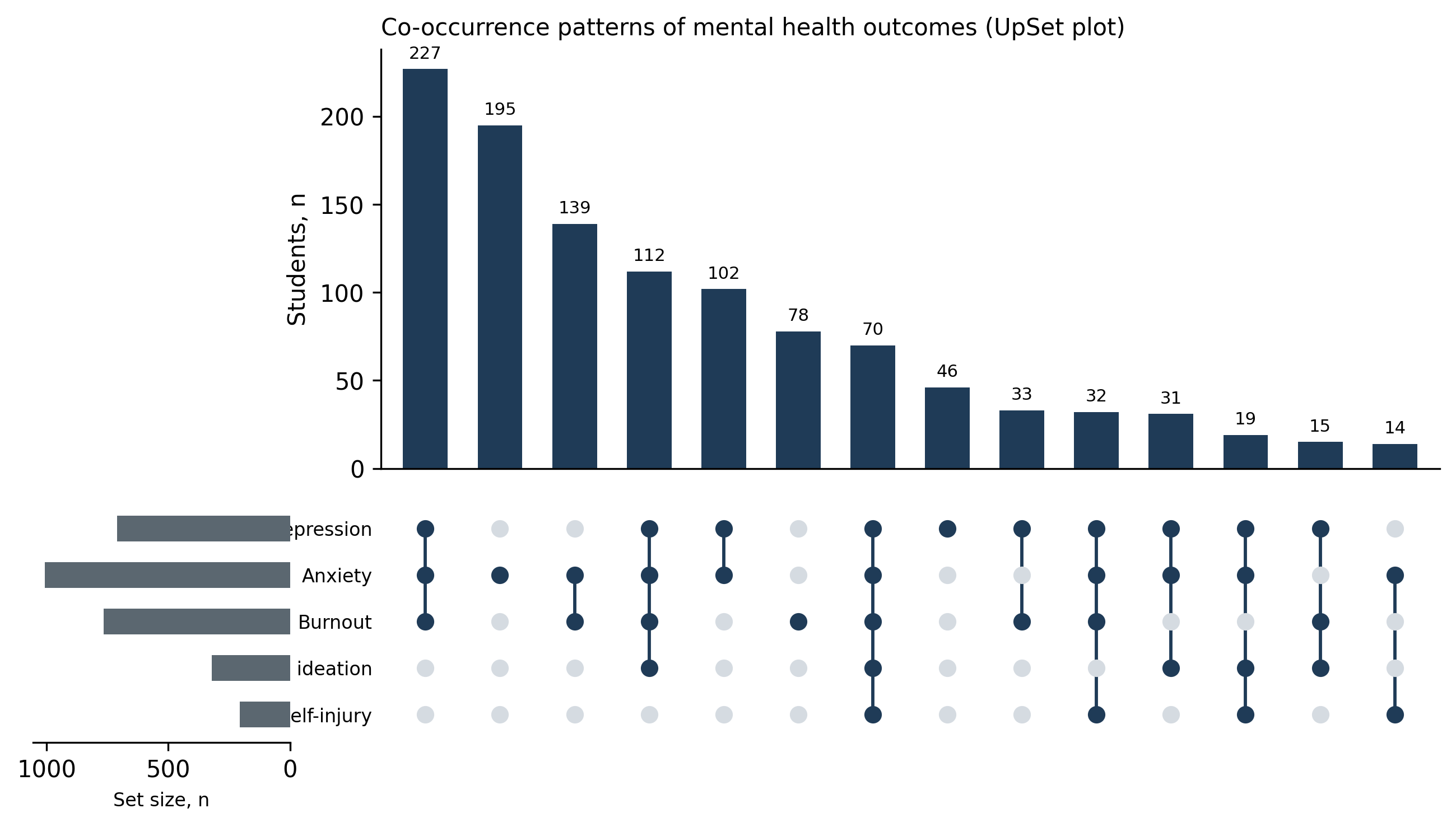

*Figure S1. Co-occurrence patterns (UpSet plot) of the five main mental health outcomes.*

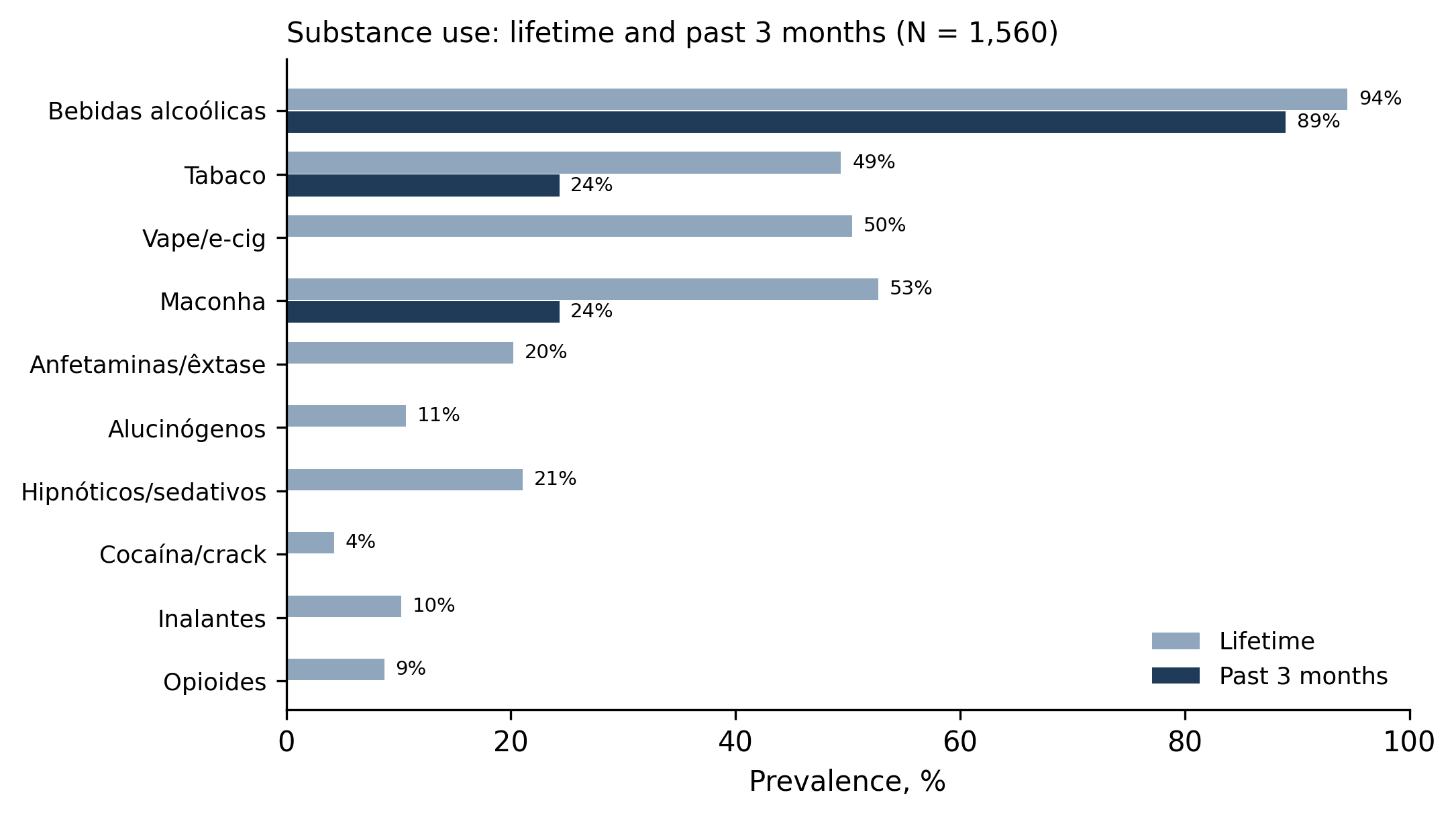

*Figure S2. Lifetime and past-3-month substance use.*

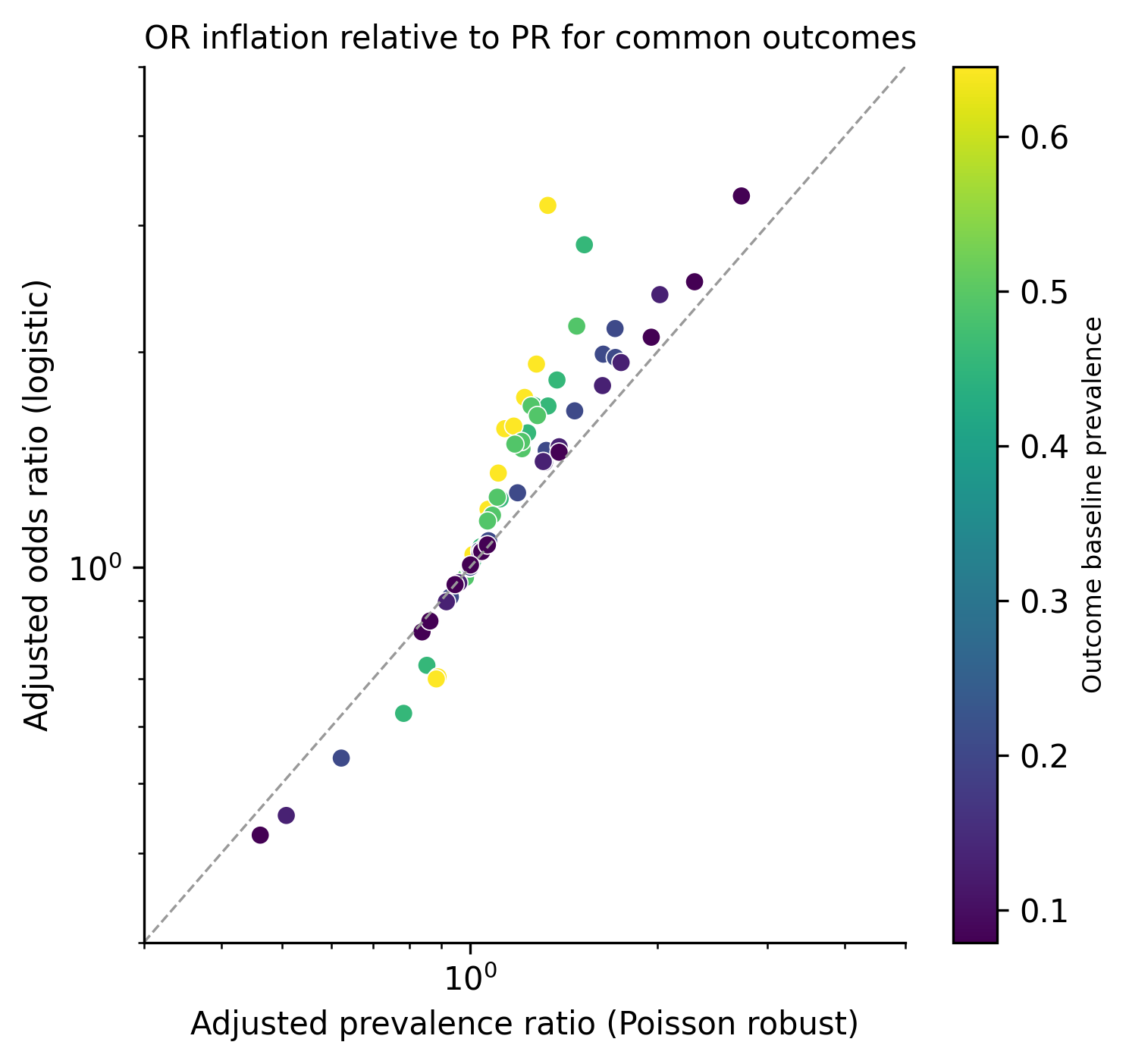

*Figure S3. Systematic inflation of adjusted odds ratios relative to prevalence ratios; colour indicates outcome baseline prevalence.*

### STROBE note

The manuscript follows the STROBE checklist for cross-sectional studies. Items covered by the present Results package: participants and flow (item 13; Results §1), descriptive data (item 14; Table 1), outcome data (item 15; Table 2, Figure 1), main results with adjusted estimates and precision (item 16; Table 3, Figure 2), other analyses — trends, interactions, sensitivity (item 17; Tables S4–S6, S8). A completed item-by-item checklist should accompany submission.
